# Quantification of abdominal fat from computed tomography using deep learning and its association with electronic health records in an academic biobank

**DOI:** 10.1101/2020.09.22.20199844

**Authors:** Matthew T. MacLean, Qasim Jehangir, Marijana Vujkovic, Yi-An Ko, Harold Litt, Arijitt Borthakur, Hersh Sagreiya, Mark Rosen, David A. Mankoff, Mitchell D. Schnall, Haochang Shou, Julio Chirinos, Scott M. Damrauer, Drew A. Torigian, Rotonya Carr, Daniel J. Rader, Walter R. Witschey

## Abstract

**Objective:** The objective was to develop a fully automated algorithm for abdominal fat segmentation and to deploy this method at scale in an academic biobank.

**Materials and Methods:** We built a fully automated image curation and labeling technique using deep learning and distributive computing to identify subcutaneous and visceral abdominal fat compartments from 52,844 CT scans in 13,502 patients in the Penn Medicine Biobank (PMBB). A classification network identified the inferior and superior borders of the abdomen, and a segmentation network differentiated visceral and subcutaneous fat. Following technical evaluation of our method, we conducted studies to validate known relationships with visceral and subcutaneous fat.

**Results:** When compared with 100 manually annotated cases, the classification network was on average within one 5 mm slice for both the superior (0.4±1.1 slices) and inferior (0.4±0.6 slices) borders. The segmentation network also demonstrated excellent performance with interclass correlation coefficients of 1.00 (p<2e-16) for subcutaneous and 1.00 (p<2e-16) for visceral fat on 100 testing cases. We performed integrative analyses of abdominal fat with the phenome extracted from the electronic health record and found highly significant associations with diabetes mellitus, hypertension, renal failure, among other phenotypes.

**Conclusion:** This work presents a fully automated and highly accurate method for the quantification of abdominal fat that can be applied to routine clinical imaging studies to fuel translational scientific discovery.

## INTRODUCTION

Medical centers collect enormous quantities of imaging data that could be extremely valuable for translational science, but many quantitative traits are not systematically extracted. One of these quantitative traits is abdominal adipose tissue volume, which is highly relevant to human health and disease and can be quantified from medical images such as CT scans. Obesity, a condition of increased adipose tissue, has been associated with numerous diseases including cardiovascular disease, diabetes, stroke, and cancer [1, 2]. However, obesity is diagnosed by body mass index (BMI) which is a poor measure of fat as it is calculated using only weight and height and does not account for variations in body composition [3].

Furthermore, not all fat is equal. Visceral adipose tissue (VAT), which is located within the abdominal cavity adjacent to vital organs, portends an even greater risk for many pathologies including cardiovascular disease, insulin resistance, and certain cancers [4]. However, the complex relationship between VAT and disease is not yet understood [4-7]. Fundamental to the continued study of this topic is the ability to accurately quantify and distinguish VAT and subcutaneous adipose tissue (SAT). Many studies rely on waist circumference (WC) as a proxy for VAT [8-10]. However, while WC has demonstrated clinical significance beyond BMI, it correlates more strongly with SAT than VAT [11, 12]. Computed tomography (CT) scans offer a solution as they are routinely performed and provide a cross sectional view of anatomy that is often used to measure abdominal adipose tissue.

Robust and automatic techniques for extracting fat imaging traits from CT could help in the at-scale task of processing data in large biobank populations, developing precision medicine algorithms, expediting clinical workflows, or even deepening our understanding of machine learning bias. There are numerous biobanks in the United States and internationally that collect genetic data and correlate this with electronic health record (EHR) documented pathology [13]. By better understanding the relationship between body composition and genetics, environment, and disease, we can begin to offer patients more targeted, precision-medicine interventions. This knowledge combined with the processing algorithm could then be integrated into a radiology practice to provide valuable information to clinicians making care decisions. Furthermore, machine learning algorithms excel in pattern recognition but sometimes make predictions that are incorrect and based on biases learned during training. [14, 15] These biases are easiest to detect when an algorithm is applied to a large diverse cohort, but this requires a method that can be efficiently applied at scale.

Many methods that have been proposed for automatic fat quantification have limitations that prevent their application to a large clinical cohort. These methods often rely on expected anatomic profiles to apply a statistical model [16-24]. The active contour model is an example of a common segmentation technique which has been applied to body composition analysis [20, 24], and works by minimizing an energy function designed to create a smooth boundary at regions of high gradients [25]. However, this approach can easily fail when image noise creates local minima or when the object has boundaries with high curvature [26]. Another approach is model based segmentation where a model is constructed based on expected geometry and then deformed to identify the object of interest on new cases [21, 23]. This approach can work well with homogenous data, but variations in shape and size limit its generalizability [27]. In summary, the above highlighted methods rely on expected attenuation profiles or geometric properties and can work well on curated datasets but easily fail when artifacts, anatomical variation, or unexpected pathology is encountered.

Deep learning offers a data-driven approach to overcome these limitations by learning and prioritizing features based on training data. Deep learning has been applied to a range of biomedical segmentation and classification tasks with impressive results [29-32]. Specific to abdominal fat quantification, deep learning has been utilized in multiple studies [28, 33-35]. However, many of these methods are only applicable to single-slice quantification and do not address identifying the slices of interest. Additionally, further work is needed to evaluate the utility of applying these deep learning approaches at scale on a diverse dataset. Altogether, these clinical and translational applications are increasingly motivating a need for automated methods to extract fat biomarkers from CT.

We built a fully automated abdomen and pelvis image curation and fat labeling technique using deep learning and applied it to CT scans to identify SAT and VAT. After technical validation, this technique was applied to 52,844 CT scans from 13,502 patients enrolled in the Penn Medicine Biobank (PMBB), a centralized resource of annotated blood and tissue samples linked with clinical EHR and genetic data. As additional validation of the methodology, we performed integrative analyses of the imaging traits with other phenotypic data extracted from the EHR including blood biomarkers, body mass index, and diagnoses (ICD-9 and ICD-10) codes.

## MATERIALS AND METHODS

### Penn Medicine Biobank

This study used data collected from participants in the Penn Medicine Biobank (PMBB). The PMBB is a resource for advanced imaging, genetics, blood biomarkers and other electronic health record data at Penn Medicine, a multi-hospital health system headquartered in Philadelphia, PA. It is a detailed long-term, prospective, epidemiological study of over 50,000 volunteers containing approximately 27,485 unique diagnostic codes (ICD-9 and ICD-10) and 1,025,963 radiology studies. All patients provided informed consent to participate in the PMBB and to utilization of electronic health record and image data, which was approved by the Institutional Review Board of the University of Pennsylvania.

### Study Cohort

At the time this study was completed (January 2020), the biobank had enrolled 52,441 patients. Participants of the PMBB were included in this study if they had an abdominal and pelvis CT scan. A detailed flowchart showing the number of participants in this study, imaging studies, and number of imaging scans (multiple scans are collected per study) is shown in Figure 1. Details of the demographic and clinical summary statistics are available in Table 1.

**Table 1.** Population characteristics for cohort

| Characteristic | Cohort (n=13,405) |  |
| --- | --- | --- |
| **Demographics** | *mean* | *std. dev.* |
| Age (years) | 57.9 | 15.2 |
| Sex | *n* | *%* |
| Male | 6,745 | 50.3 |
| Female | 6,653 | 49.7 |
| Ancestry |  |  |
| European | 8,400 | 62.7 |
| African | 3,490 | 26.0 |
| Asian | 213 | 1.6 |
| Other/Unknown | 1302 | 9.7 |
| **Clinical Metrics** | *mean* | *std. dev.* |
| Height (m) | 1.70 | 0.10 |
| Weight (kg) | 85.6 | 22.1 |
| Body mass index, kg/m<sup>2</sup> | 28.8 | 7.0 |
| Systolic Blood Pressure, mmHg | 126.5 | 12.0 |
| Diastolic Blood Pressure, mmHg | 74.8 | 7.3 |
| **Diagnoses** | *N* | *%* |
| Hypertension |  |  |
| Yes | 7,103 | 63.3 |
| No | 5,409 | 36.7 |
| Diabetes |  |  |
| Yes | 3,650 | 35.0 |
| No | 8,862 | 65.0 |
| Heart Failure |  |  |
| Yes | 2,497 | 22.6 |
| No | 10,015 | 77.4 |
| Ischemic Heart Disease |  |  |
| Yes | 3,109 | 27.1 |
| No | 9,403 | 72.9 |
| Renal Failure |  |  |
| Yes | 3,672 | 35.5 |
| No | 8,840 | 64.5 |
| **Lab Values** | *mean* | *std. dev.* |
| Cholesterol, mg/dL | 176.0 | 39.1 |
| HDL cholesterol, mg/dL | 50.8 | 15.5 |
| LDL cholesterol, mg/dL | 98.6 | 32.2 |
| Triglycerides, mg/dL | 124.8 | 64.2 |
| HbA1c, % | 6.4 | 2.5 |
| BUN, mg/dL | 17.9 | 9.8 |
| Creatinine, mg/dL | 1.01 | 0.42 |

**Figure 1:**
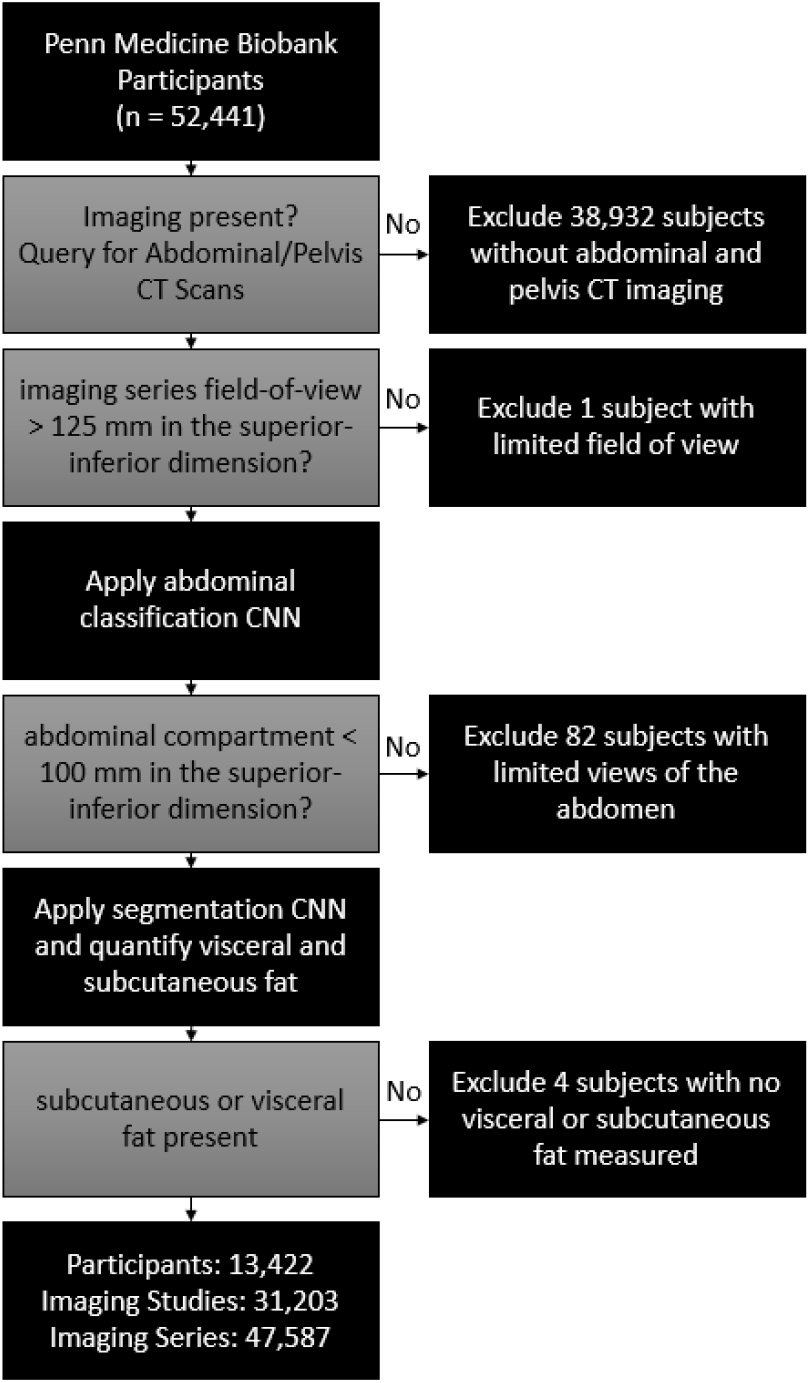
Detailed description of the Penn Medicine Biobank participants included in this study and exclusion criteria applied at each step of analysis. An imaging study was defined as a single visit and one or more CT scans of the abdomen or pelvis. An imaging scan was defined as a set of CT images with or without contrast; multiple scans were performed per imaging study. Abdominal classification CNN is the network used to identify axial CT views that show the abdomen. Segmentation CNN is the network which delineates the abdominal contour. After applying exclusion criteria, association studies were performed using body mass index, blood biomarkers and diagnostic codes.

### Image Analysis

A schematic of the overall approach to subcutaneous and visceral fat segmentation using deep learning is shown in Figure 2. Two deep learning neural networks were used to 1) classify 2D images of the abdomen or pelvis as showing the abdomen and then 2) segment these images for the abdominal compartment. In a final step, intensity-based thresholding criteria were applied to visceral and subcutaneous compartments to label fat in these regions. Processing was performed by utilizing distributive cloud computing across 50 virtual instances each equipped with an NVIDIA K80 GPU. Please see supplement for further details on our data, model training, and prediction framework.

**Figure 2:**
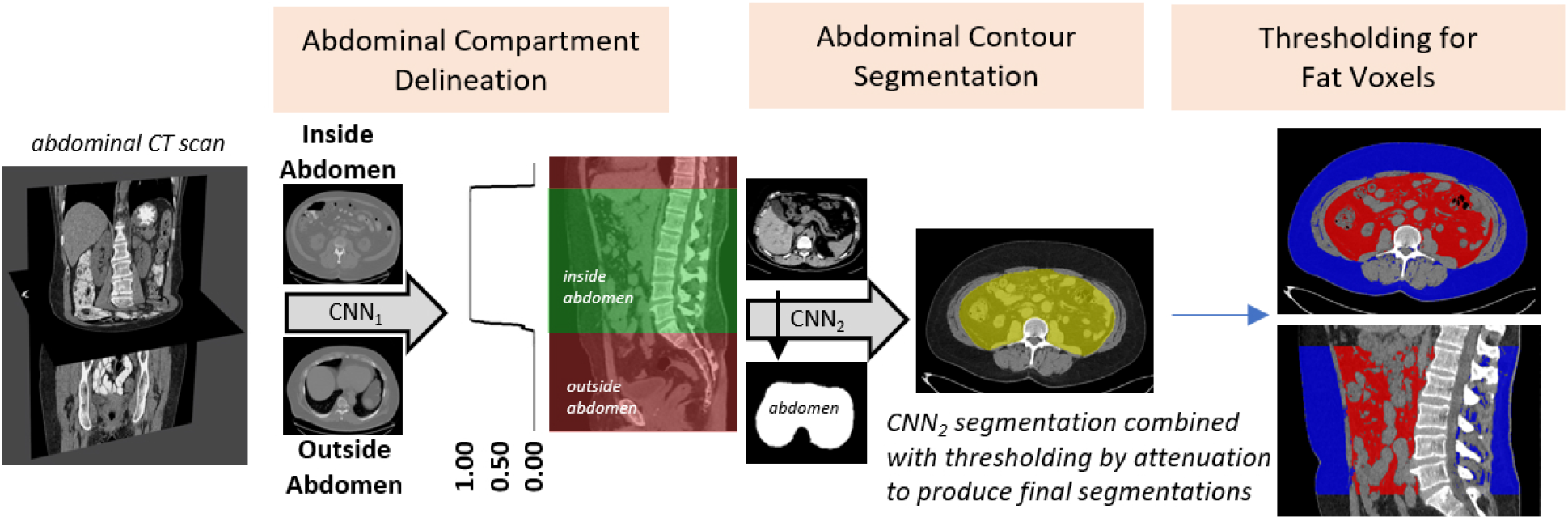
Automated extraction of abdominal fat from CT scans. Abdominal classification network (CNN_1_) classifies 2D image slices as belonging inside the abdomen or outside the abdomen. Segmentation network (CNN_2_) delineates the border of the inner abdominal contour. Fat voxels are identified based on CT attenuation and those within the contour represent visceral and those outside represent subcutaneous adipose tissue. CT=Computed Tomography, CNN=Convolutional Neural Network.

#### Classification Model – Identification of Images Showing Abdominal Anatomy

The first network labeled 2D slices as belonging to the abdominal cavity. Four candidate deep learning architectures were evaluated for this task, selected based on excellent performance demonstrated in the literature for classification tasks: VGG-16 [39], ResNet-50 [40], Inception V3 [41], and DenseNet-121 [42]. These architectures were trained once with randomly initialized weights and then again using pre-trained weights from the ImageNet dataset for a total of 8 model variants. The plus sign (eg. VGG-16+) will be used to indicate an architecture trained using ImageNet weights. The networks were trained to output a probability between 0 and 1 indicating the likelihood that the slice is within the boundary of the abdomen. To extract the boundaries from this array of probabilities we first subtracted 0.50 from every value such that such that values were in the range [-0.5, 0.5]. Next, we applied Kadane’s algorithm to find the contiguous sub-list of these values that gives the maximum sum [42].

Training was conducted on a set of 468 scans of which 375 scans (35,305 slices) were used for the training set and 93 scans (8,775 slices) for the validation set. Performance of all 8 networks (4 distinct architectures, trained with and without pretrained weights) was then evaluated for its sensitivity, specificity, and accuracy on a separate testing set of 100 scans which were randomly selected from the PMBB. For each network, these metrics were first calculated individually on each scan and then the metrics were averaged across all 100 scans. For both the classification and segmentation tasks, metrics were compared using pairwise t-tests with Bonferroni multiple comparison correction and a level of significance of 0.05. The highest performing network was selected, and 5-fold cross validation was then performed on this network using all available 568 scans.

#### Segmentation Model Architecture – Labeling of subcutaneous and visceral fat pixels

The second network delineated SAT and VAT from axial 2D slices. Three candidate deep learning architectures were selected based on excellent performance in the literature for segmentation tasks: U-Net [44], Deep Lab V3 using Xception encoder [45, 46], and Deep Lab V3 using MobileNet V2 encoder [45, 47]. The networks output a probability for each voxel indicating the probability that it belongs to the foreground, and probabilities greater than or equal to 0.50 are attributed to the foreground. Additional post-processing steps include only preserving the largest connected component and filling any holes for each slice.

Training was conducted on a set of 62 scans with 50 scans (2,059 slices) randomly selected for the training set and 12 scans (498 slices) for the validation set. Performance of these networks was evaluated on a separate testing set of 20 scans which were randomly selected from the PMBB. Region-of-interest area overlap ratios (Dice scores [48]) were calculated to measure agreement between manual and automatic segmentations for the abdominal contour as well as SAT and VAT. The highest performing network was selected, and then 5-fold cross validation was conducted on this network using all 82 scans. For additional evaluation, 100 abdomen/pelvis CT scans were randomly selected from the PMBB, and VAT and SAT was manually segmented on a single slice between L3 and L4. These values were compared to automatically derived measurements from the highest performing model.

### Association Studies

#### BMI Correlation Analysis

The highest performing networks for both the classification and segmentation tasks were then used to process all 31,419 studies. The CNN derived metrics for SAT and VAT were further validated by comparing these values to clinically assessed BMI values. The most recent BMI measurement was attributed to each scan, and measurements acquired greater than 365 days from the scan were excluded. Pearson’s correlation coefficient was computed to measure the degree of association. In all association analyses, we utilized the average metric area of SAT or VAT across all slices. To associate a single fat value to each patient, we took the median (or mean if exactly two) value for studies with multiple scans or for patients with multiple studies. This approach for attributing a single image derived phenotype to a patient was utilized in all association studies.

#### Phenome-wide Association Study

A phenome-wide association study (PheWAS) was performed to investigate the phenotypic associations of having a higher VAT-SAT ratio (VSR). ICD10 codes were first mapped to ICD9 codes using the 2017 general equivalency mapping (GEM). Next, ICD9 codes were aggregated into phecodes using the PheWAS R package to create 1,816 phecodes. Patients with at least 2 occurrences of a phecode are considered cases, those with none are controls, and those with 1 are treated as missing. Phecodes with less than 100 cases were excluded. Logistic regression was then performed with each phecode as the outcome and VSR as a predictor. Regression was performed controlling for the covariates of age, sex, and race. Bonferroni multiple comparison correction was used to determine the level of significance.

#### Relationship between Lab Values and VSR

To investigate the relationship between VSR and common clinical laboratory studies, VSR values were organized into quartiles. Since VSR is associated with both sex and race, patients were first stratified into four categories based on sex and race. Patients with the highest and lowest 25% of values within each of these categories were identified and the distribution of laboratory measures between the high and low groups was compared. Lab values acquired greater than 90 days and BMI measurements acquired greater than 365 days from the scan-date were excluded. To compare the distributions, a Wilcox rank-sum test was performed.

## RESULTS

### Patient Cohort Acquisition and Analysis

After applying exclusion criteria, 31,419 studies containing a total of 52,844 scans corresponding to 13,502 patients were analyzed to identify SAT and VAT. Processing was performed in 5 hours and 8 minutes (0.35 seconds/scan) by using parallel processing. The total runtime across all 50 instances was 201 hours and 43 minutes (13.7 seconds/scan). Following the exclusion of additional scans based on the size of abdominal window and volume of fat detected, 31,158 studies containing 47,470 scans corresponding to 13,405 patients were utilized in association studies (Figure 1).

### Image Analysis – Technical Validation

#### Localizer Network

Without the use of pretrained weights, the VGG-16, ResNet-50, Inception V3, and DenseNet-121 networks converged after 27, 59, 54, and 40 epochs, respectively. When using ImageNet weights, the networks converged more quickly after 9, 17, 8, and 17 epochs, respectively. The performance of the classifier networks is shown in Table 2. All architectures achieved a sensitivity greater than 0.97, specificity greater than 0.98, and accuracy greater than 0.98. Using pre-trained weights for model training did not significantly increase sensitivity for any of the architectures (p≥0.081), but specificity (p=1.4E-5) and accuracy (p=3.9E-7) did increase for ResNet-50. When comparing the metrics between all architectures, VGG-16+ had a significantly greater average sensitivity (p=0.025), specificity (p=5.9E-5), and accuracy (p=2.5E-7) than ResNet-50 but pairwise t-tests between VGG-16+ and the other architectures demonstrated no difference (p≥0.18). Regarding runtime, the VGG-16, ResNet-50, Inception V3, and DenseNet-121 models took on average 4.2, 3.6, 5.0, and 4.3 seconds, respectively, per scan to process the 100 testing cases.

**Table 2.** Performance metrics for classifier networks. 1^st^ column indicates the type of classifier architecture with plus-signs for architectures that used pretrained weights from ImageNet. The last entry in this column (VGG-16+ 5-Fold) is for the 5-Fold cross validation performed on the VGG-16+ architecture. 2^nd^ and 3^rd^ columns list the mean±standard deviation of the number of slices the predicted border is from the ground truth border for the inferior and superior boundary. 4^th^ through 6^th^ column list the mean±standard deviation for sensitivity, specificity, and accuracy in classifying the 2D slices.

|  | **Inferior** | **Superior** | **Sensitivity** | **Specificity** | **Accuracy** |
| --- | --- | --- | --- | --- | --- |
| **VGG-16** | 0.40±0.65 | 0.55±1.40 | 0.988±0.031 | 0.991±0.015 | 0.990±0.014 |
| **DenseNet-121** | 0.61±1.05 | 0.67±1.32 | 0.978±0.042 | 0.990±0.012 | 0.986±0.017 |
| **ResNet-50** | 0.82±1.11 | 1.03±1.76 | 0.972±0.053 | 0.982±0.024 | 0.978±0.022 |
| **Inception V3** | 0.42±0.64 | 0.38±1.09 | 0.986±0.034 | 0.987±0.017 | 0.986±0.016 |
| **VGG-16+** | 0.42±0.64 | 0.38±1.09 | 0.989±0.025 | 0.993±0.014 | 0.991±0.013 |
| **DenseNet-121+** | 0.49±1.24 | 0.30±1.03 | 0.987±0.032 | 0.995±0.010 | 0.992±0.014 |
| **ResNet-50+** | 0.36±0.73 | 0.41±1.02 | 0.988±0.030 | 0.993±0.013 | 0.991±0.013 |
| **Inception V3+** | 0.42±0.56 | 0.54±1.37 | 0.985±0.035 | 0.993±0.011 | 0.990±0.015 |
| **VGG-16+ 5-Fold** | 0.32±0.88 | 0.41±0.98 | 0.988±0.029 | 0.994±0.013 | 0.992±0.014 |

VGG-16+ was selected as the architecture of choice based on its non-inferior performance, simplicity of design, and fast runtime. The automated method was on average within one 5 mm slice from the manually selected slice for both the superior (0.4±1.1 slices) and inferior (0.4±0.6 slices) borders. For the superior border, the prediction ranged from 3 slices below to 3 slices above the manual label. For the inferior border, they ranged from 6 slices below to 8 slices above (one vertebral level) the manual label. Before application of the maximum sub-list algorithm, the VGG-16+ algorithm had a sensitivity of 0.99±0.03, specificity of 0.99±0.01, and accuracy of 0.99±0.01. These metrics were without significant change after application of Kadane’s algorithm – p-values of 0.82, 0.65, 0.92, respectively. 5-fold cross validation on the VGG-16+ architecture was then performed using all available 568 studies. It demonstrated excellent sensitivity, specificity, and accuracy values with average values of 0.99 for all three metrics (Table 2). These metrics obtained during cross validation were not significantly different from those obtained with VGG-16+ on the testing set (p≥0.32).

#### Segmentation Network

The U-Net, DeepLab+MobileNet V2 and DeepLab+Xception converged after 39, 70, and 63 epochs, respectively. When assessing performance on the testing set of 20 scans, all architectures achieved mean Dice values greater than or equal to 0.98 for the abdominal contour as well as the SAT and VAT regions (Table 3). When conducting pairwise comparison between the three algorithms, there was no significant difference in the means for any of the metrics (p≥0.16). Regarding runtime, U-Net, DeepLab+MobileNet V2 and DeepLab+Xception ran in 7.3, 7.3, and 8.5 seconds per case, respectively. The U-Net architecture was selected for its non-inferior performance, runtime efficiency, and simplicity of design. 5-fold cross validation was then performed for the U-Net architecture, and it obtained mean Dice values of at least 0.97 for all three metrics (Table 3). The metrics obtained during cross validation were also not significantly different from those obtained with U-Net on the original testing set (p≥0.11).

**Table 3.** Performance metrics for segmentation networks. 1^st^ column indicates type of segmentation architecture. The ‘U-Net 5-fold’ row provides values for the 5-fold cross validation performed on the U-Net architecture. The ‘U-Net Extended Training’ row is evaluation for predictions using weights when the U-Net was run for an extended duration past the normal stopping criteria. The 2^nd^ through 4^th^ columns provide mean±standard deviation Dice values for the abdominal contour, subcutaneous fat, and visceral fat when comparing manual to automated predictions.

|  | **Abdominal Contour** | **Subcutaneous Fat** | **Visceral Fat** |
| --- | --- | --- | --- |
| **U-Net** | 0.980±0.008 | 0.998±0.003 | 0.991±0.007 |
| **DeepLab+MobileNet V2** | 0.975±0.008 | 0.998±0.002 | 0.986±0.009 |
| **DeepLab+Xception** | 0.979±0.006 | 0.998±0.001 | 0.990±0.006 |
| **U-Net 5-fold** | 0.972±0.052 | 0.978±0.072 | 0.997±0.008 |
| **U-Net Extended Training** | 0.982±0.007 | 0.998±0.002 | 0.992±0.006 |

In our experience, while extending the duration of training does not yield a significant improvement in Dice metrics it can result in better performance on edge cases, provided the model is not allowed to overfit. For this reason, we trained the U-Net for 345 epochs and selected epoch 326 based on Dice values for the validation set; these weights were used for processing studies at scale. Dice metrics for evaluation of this model are shown in Table 3 and representative segmentation results are shown in Figure 3. Scatterplots and Bland-Altman plots showing evaluation results for the single slice segmentations on 100 different patients is shown in Figure 4 (Panels A-D). Interclass correlation coefficients of 0.9999 (p<2e-16) and 0.9998 (p<2e-16) were achieved for the prediction of SAT and VAT, respectively. There was a significant bias of 1.4±1.6 cm^2^ (p=3.1e-15) for subcutaneous and -1.4±1.6 cm^2^ (p=3.1e-15) for VAT. For this analysis, the average areas were 298.7±167.1 cm^2^ for subcutaneous and 153.5±109.6 cm^2^ for VAT.

**Figure 3:**
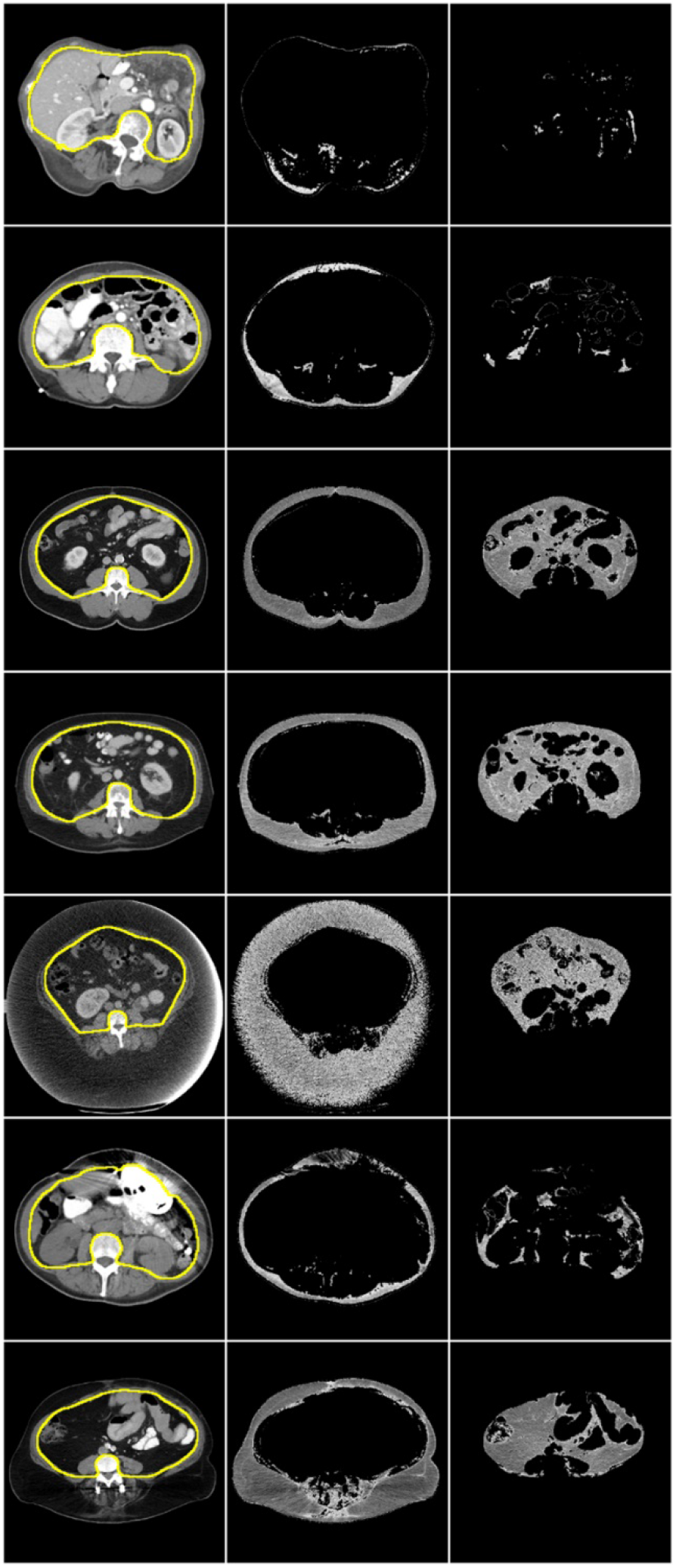
Representative segmentation results from six different patients. First column shows CT slice with yellow line indicating the CNN predicted contour. From this contour, our algorithm identifies subcutaneous fat (second column) and visceral fat (third column). Row 5 shows a patient with beam hardening artifact from a left ventricular assist device, and row 6 shows a patient with subcutaneous scar tissue from a spinal fusion surgery.

**Figure 4:**
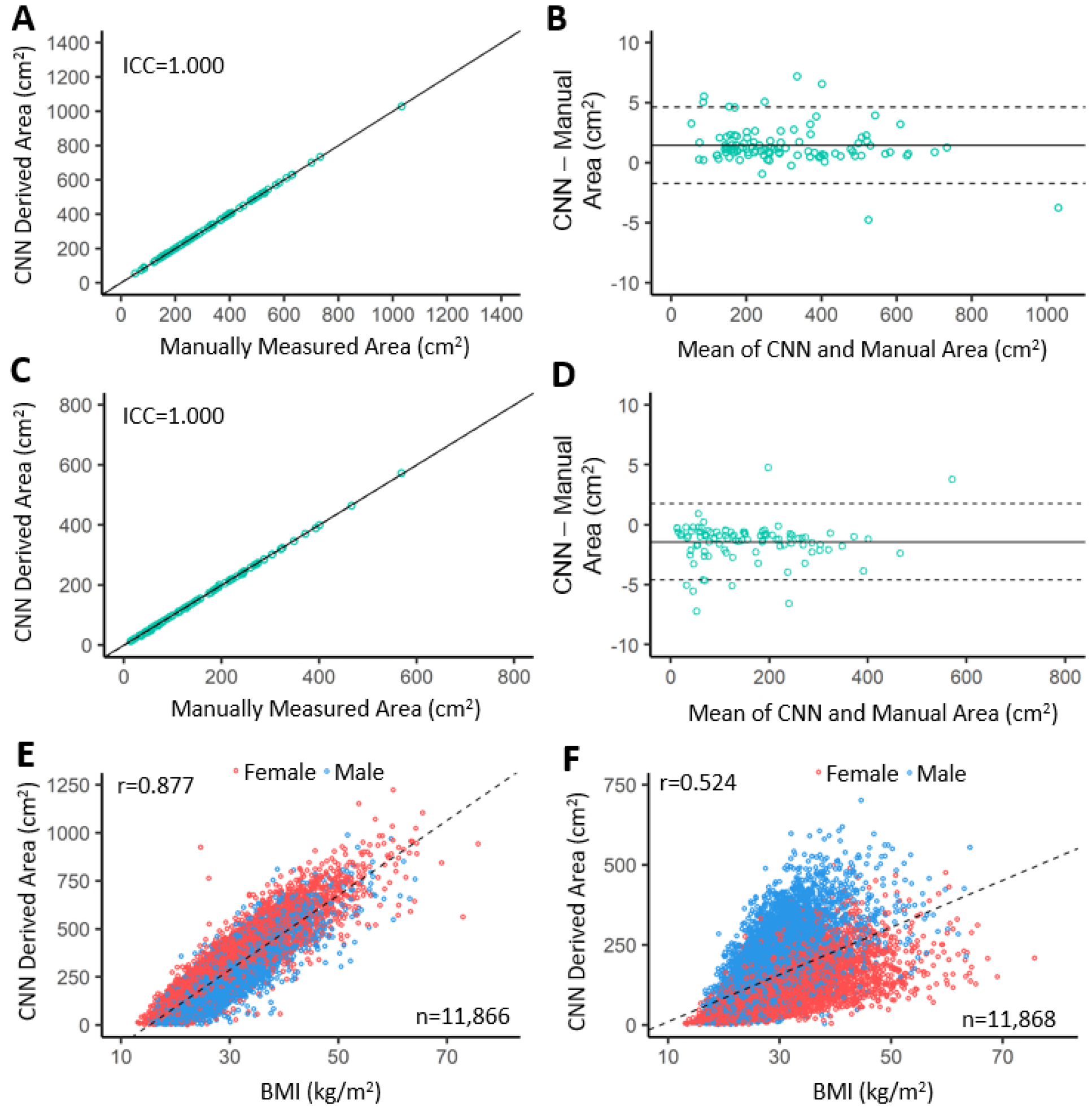
Scatterplots and Bland-Altman plots comparing CNN to manually derived area for subcutaneous (A, B) and visceral (C, D) fat where fat was derived from a single slice on 100 randomly selected scans. Panels E and F show scatterplots of CNN derived area versus BMI for subcutaneous (E) and visceral (F) fat where fat was derived from all available biobank scans. CNN=Convolutional Neural Network, ICC=Interclass Correlation Coefficient, BMI=Body Mass Index.

### Association Studies

Association studies were conducted investigating the relationship between the CNN derived fat values and BMI, clinical lab values, and billing codes. The relationships between the CNN derived fat values and BMI are shown in Figure 4 (Panels E and F). There was a significant correlation between BMI and both subcutaneous (r=0.876, p<2e-16) and visceral (r=0.522, p<2e-16) fat.

Next, we compared the distribution of lab values and BMI measurements for patients in the bottom quartile of VSR values to those in the top quartile. For the high VSR group there was a significant increase in triglycerides (p=5.9e-10), glycated hemoglobin (A1C; p=2.0e-4), blood urea nitrogen (BUN; p=1.0e-14), creatinine (p=9.8e-55) and BMI (p=1.7e-5). There was also a significant decrease in high-density lipoprotein (HDL; p=0.0014). Density plots showing the distribution of values for the two groups are shown in Figure 5.

**Figure 5:**
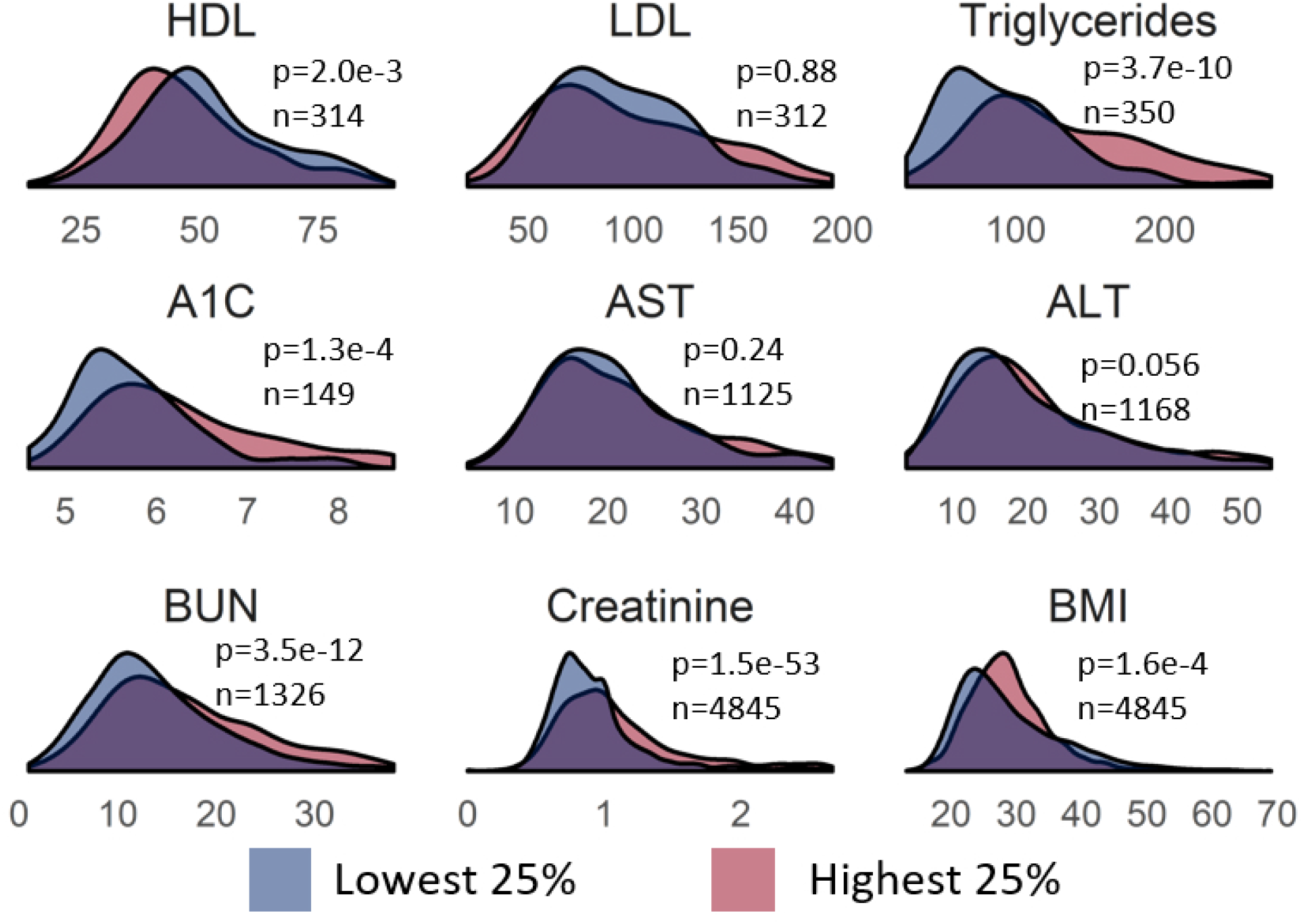
Biomarker distributions dichotomized by VSR values. HDL = High density lipoprotein, LDL = low density lipoprotein, A1C = glycated hemoglobin, AST = aspartate aminotransferase, ALT = alanine aminotransferase, BUN = blood urea nitrogen, BMI = body mass index.

A phenome-wide association study of VSR revealed significant associations with several pathologies. The plot is shown in Figure 6. The strongest association was with diabetes mellitus (p=1.7e-23). There were multiple hits for other endocrine disorders related to diabetes or lipid metabolism. Within the circulatory system, the strongest signals were for hypertension (p=2.5e-19) and hypertension-related complications. In the renal system, there were seven significant hits, including chronic kidney disease (p=5.4e-18), renal failure (p=1.7e-15), and renal transplant (p=1.6e-12).

**Figure 6:**
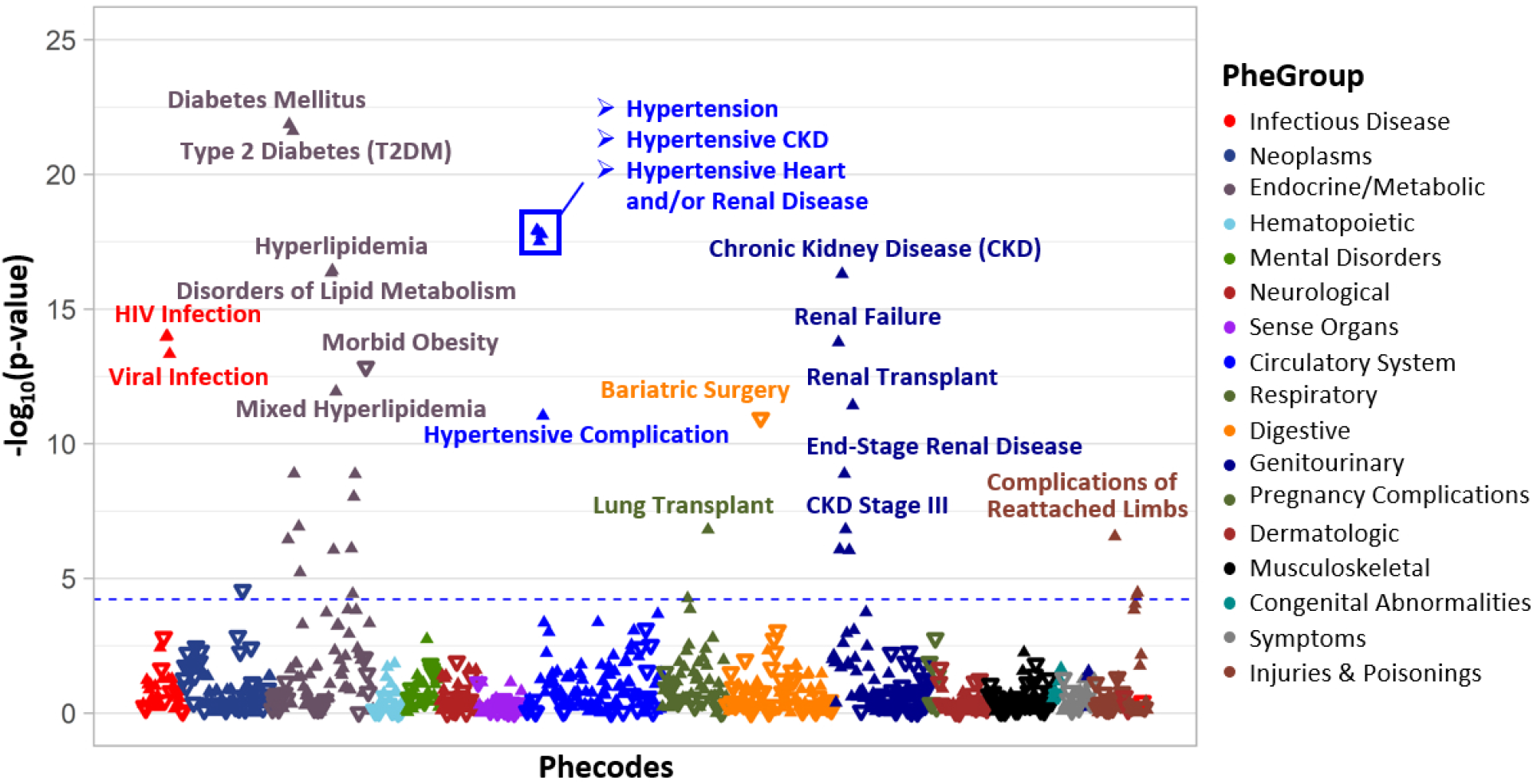
Phenome-Wide Association Study of VSR. Blue line indicates level of significance with Bonferroni multiple-comparison correction. Upward-facing triangles indicate a positive association and downward-facing triangles indicates a negative association.

## Discussion

In this paper, we present a fully automated approach to accurately quantify abdominal fat from clinical CT scans. We provide a rigorous evaluation of several prominent deep learning architectures for this task as well as an evaluation on the use of transfer learning by using ImageNet weights. Following architecture selection, we utilized distributive processing in a cloud computing environment to quantify full abdominal volumes of visceral and subcutaneous fat from 52,844 scans in 5 hours (∼172 scans per minute). This demonstrates the efficiency of a fully trained deep learning network to label abdominal CT images at a high rate which is necessary to perform large-scale research applications or to support automatic, quantitative reporting of abdominal fat by radiologists. To our knowledge, this is the first automatic technique for 3D CT abdominal fat segmentation to be associated at scale to unbiased EHR data from an academic biobank.

During the technical validation of our methods, the results showed outstanding agreement between manual and automated measurements of fat. There was a significant bias of 1.4 cm^2^ for both SAT and VAT. However, compared to the average areas of 299 cm^2^ for SAT and 154 cm^2^ for VAT in our testing set, this bias represented a small error (<1%). While previous studies that utilized radial projections or shape analysis were not tested on patients with extreme body habitus [16, 54], our method demonstrated excellent performance on both the randomly selected testing set and during 5-fold validation which included testing on many studies which were intentionally selected for either noise or extreme body habitus. Figure 3 highlights the fault-tolerance of the algorithm to variations in patient body habitus and BMI. Indeed, when representative diverse cases are included in the training data, data-driven segmentation correctly quantified fat area when there was no visible SAT or when body habitus or post-surgical changes caused significant artifacts. Work by Weston et al. applied a similar U-Net CNN for body composition analysis to manual selected CT slices and achieved Dice scores of 0.98±0.03 for SAT and 0.94±0.12 for VAT [33]. These are comparable to our values of 0.998±0.002 for SAT and 0.992±0.006 for VAT. Given that we used a similar method, the small improvement in our approach is likely due to the diversity of pathology in our training data.

We found that the association between SAT or VAT and BMI was strong and similar compared with previous studies that found associations of r=0.73-0.93 for SAT and r=0.61-0.77 for VAT [7, 55, 56]. In agreement with these studies, we found a stronger association between BMI and SAT than between BMI and VAT. When comparing lab values between patients with high and low values of VSR, the high VSR group had significantly higher triglycerides and lower HDL but no significant change in LDL. This was consistent with previous findings that VSR was positively associated with triglycerides, negatively associated with HDL, and showed no significant association with LDL [19]. Glycated hemoglobin (A1C) levels were significantly elevated in our analysis which aligns with a known association with diabetes [4]. While an association between VSR and liver enzymes has been reported [57], our analysis did not find significant associations for AST or ALT. Our finding of increased creatinine and BUN in the higher VSR group is consistent with previous findings that increased VAT is associated with decreased renal function and progression to end-stage renal disease, particularly in diabetic kidney disease [58].

By investigating association between VSR and phecodes, we were able to interrogate disease associations in an unbiased manner. As expected, we found numerous associations with diabetic, hypertensive, and kidney disease pathologies. There were some unexpected associations with transplants and human immunodeficiency virus (HIV) infections. The association with HIV is likely due to lipodystrophy which is commonly seen in these patients [59]. Similarly, the association with transplants is likely secondary to the use of corticosteroids following transplant which is known at high doses to increase the amount of VAT [60]. The negative association with bariatric surgery is likely because patients who get bariatric surgery have significant depots of subcutaneous fat.

While the VGG-16 and U-Net networks are frequently applied for medical imaging applications, there are multiple technical and translational advances demonstrated in this paper. These major contributions include, 1) developing a two-step classification-segmentation pipeline that efficiently processes scans without the need for any manual input, 2) providing rigorous comparisons of multiple deep learning architectures for this application and 3) conducting association studies between image features and phenotypes in an academic biobank to provide both additional validation as well as to highlight the utility of applying our method at scale.

There are several limitations to this study. While the attenuation range for fat has been defined in literature, CT scans acquired over several decades may contain artifacts or utilize reconstruction algorithms that distort attenuation. Specifically, implanted devices can distort attenuation by creating beam hardening artifacts or scar tissue can be introduced changing the tissue attenuation by physiologic means. Given our automated approach, it is also possible that scans do not reach the inferior or superior borders of the abdomen, and this could skew the resulting fat values. This study also derived disease phenotypes from EHR billing codes which are often incomplete. As this was a retrospective cohort, there could be significant selection bias for sicker patients or certain diseases based on biobank recruitment methods. Further work should be performed to investigate the associated phenotypes to refine our understanding and identify any causative relationships. Additionally, there is great potential in the utilization of this method in the context of a biobank such as the PMBB where genetic information is available. This may provide greater insight into the mechanism of pathogenicity for VAT, which would be of great interest to the scientific community.

In conclusion, this study presents a fully automated method for the quantification of abdominal fat that functions with high accuracy and can be applied efficiently in a cloud computing environment. This method has been validated through traditional technical approaches and by integration of our results with clinical data. This integration further highlights how autonomous image trait quantification can facilitate translational research especially in the context of an academic biobank.

## Data Availability

Data is available upon reasonable request to the authors.

## Contributorship Statement

### Author contributions

W.R.W., D.R., R.C., and M.M. obtained the funding. W.R.W., D.R., R.C., and M.M. were responsible for the concept and design of the study. W.R.W., D.R., M.M., M.S., D.M., and A.B. were involved in patient identification and data procurement from the clinical workflow. M.M., Q.J., J.C., D.T., and M.R. were involved in the process of annotating ground-truth data. W.R.W., M.M., H.S. and H.L. were involved in model training and evaluation. W.R.W., M.M., H.S., M.V., Y.K., and S.D. were involved in the statistical analysis. W.R.W., D.R. and M.M. drafted the manuscript, and all authors revised and approved the final manuscript.

## Data availability statement

Data is available upon reasonable request to investigators.

## Additional methods

### Study Cohort

Participants of the PMBB were included in this study if they had an abdominal and pelvis scan as indicated by an Current Procedural Terminology (CPT) code 74176 (CT abdomen and pelvis without contrast), 74177 (CT abdomen and pelvis with contrast), and 74178 (CT abdomen and pelvis with and without contrast). There were respectively 10,086, 19,415, and 1,923 studies for these CPT codes. <u>_[MM1]_</u>Imaging scans were excluded if they were less than 125 millimeters (mm) in the superior-inferior dimension, as they would be unlikely to include the entire abdominal field; this dimension was calculated by multiplying the number of slices in the scans and the slice thickness. Scans were also excluded if the abdominal compartment was determined to be less than 100 mm in the superior-inferior dimension; this dimension was calculated by multiplying the number of slices in the abdominal compartment, as determined by the classification network, and the slice thickness_.<u>[MM2]</u>_

### Image Analysis

#### Ground truth data

Ground truth data was provided to the classification and segmentation networks for training. All training data was manually generated by a trained technician under the supervision of a board-certified abdominal radiologist. All manually labeled scans were 5 mm slice thickness. Since the algorithm operated on 2D image slices, it was easily extended to scans of variable thickness. The breakdown of scan characteristics including slice thickness, number of slices, and abdominal compartment height is shown in Supplementary Table 1 for the entire cohort. _[MM3]_The two-category classification network identified axial CT images located in the abdomen (e.g. 0 = not in abdomen, 1 in abdomen). The superior border of the abdomen was defined as the first axial view that the lungs were no longer visible (when moving inferiorly). The inferior border was defined as the first axial view that the bottom of the L5 vertebra was visible (when moving superiorly). These borders were manually labeled on 468 training scans of which 375 scans (35,305 slices) were used for the training set and 93 scans (8,775) for the validation set – an 80:2split. A separate testing set of 100 scans was randomly selected from available studies and manually labeled. When 5-fold cross validation was performed, it utilized all 568 annotated scans. In each fold, of the scans allotted for model training, we used the same 80:20 split for training and validation sets and all the same configurations as previously described.

The segmentation network was trained to label pixels inside the abdomen. The network was trained on a total of 62 scans with 50 scans (2,059 slices) randomly selected for training and 12 scans (498 slices) for validation – an 80:20 split. Training data was selected iteratively when the model underperformed on a scan. A separate testing set of 20 scans was randomly selected and segmented. When performing 5-fold cross validation, all 82 segmented scans were utilized. Of note, the network was not trained to directly label subcutaneous and visceral fat. Rather, the segmentation consisted of two steps, where 1) the abdominal compartment was segmented, as shown in Figure 2, and 2) fat voxels were located by thresholding for HU between -190 and -30 [1, 2]. _[MM4]_The location of the abdominal compartment segmentation was chosen based on the definitions of subcutaneous and visceral fat by Shen et al. [3], where the definition of subcutaneous fat includes fat located between muscles._[MM5]_ All fat voxels within the abdominal compartment segmentation are visceral fat and all those outside the segmentation are subcutaneous fat. While it would be possible to directly predict fat voxel type using machine learning, the attenuation range for fat voxels is well defined, and we can simplify the learning task by training the model to identify the abdominal compartment instead of identifying the fat voxels directly_.[MM6]_ Image augmentation included variations in zoom, rotation, horizontal flipping, brightness, as well as width and height shifting. Image shear was also utilized to augment the dataset by providing minor distortions in patient anatomy. _[MM7]_

#### Hardware Specifications and Data Processing

All architectures were implemented in Python using the Tensorflow package. Training was conducted using an NVIDIA P100 graphical processing unit (GPU) and inference utilized NVIDIA K80 GPUs. The network inputs were 2D axial slices with size of 256×256 pixels. A window width of 1800 HU and level of 400 HU was used for the abdominal compartment delineation network and a window width of 400 HU and a level of 50 HU was used for the abdominal contour segmentation network. These window width and level values were used both for the network inputs as well as during manual segmentation. _[MM8]_

Processing was performed by utilizing distributive cloud computing across 50 virtual instances each equipped with an NVIDIA K80 GPU. All scans were stored in a cloud bucket located in the same geographic region as the instances. Before launching the instances, a local machine queried the bucket to identify all scans for processing. This list was divided into 50 equal sublists and each instance was automatically launched and responsible for processing one of the sublists. In this way, we eliminated the risk of duplicate processing or need for communication between instances.

#### Classification Model – Identification of Images Showing Abdominal Anatomy

Four candidate architectures were evaluated for the classification task: VGG-16, ResNet-50, Inception V3, and DenseNet-121. Image inputs were resized to be compatible with the architectures – (224, 224, 3) for VGG-16, ResNet-50, and DenseNet-121 and (299, 299, 3) for Inception V3. The three image channels were obtained simply by stacking each 2D slice upon itself. This was necessary to use the native configuration of these models. Training was stopped when the network advanced 10 epochs without an improvement of at least 0.0005 in average class accuracy when assessed on the validation set; the epoch with maximum average accuracy was then selected.

With regards to the VGG-16 architecture used in our paper (Supplementary Figure 1), the key differences with the implementation originally described in Simonyan for the ImageNet 2014 challenge were the following:

1. The proposed network may best resemble the network B described in [4], which has two convolutional layers before pooling each layer. There were three fully connected intermediate layers in each. In our network, the final fully connected output layer had two output categories instead of 1000 categories used for image classification; this layer was activated using a softmax function.
2. There were differences in the input image matrix size and number of filter channels per layer. These differences were mainly to accommodate the larger image size of the CT images.
3. Pooling and upsampling steps were similar in our network. Rectified Linear Units (ReLu) were used in all convolutional layers for activation.

#### Segmentation Model Architecture – Labeling of subcutaneous and visceral fat pixels

Three candidate architectures were evaluated for the segmentation task: U-Net, Deep Lab V3 with Xception encoder and Deep Lab V3 with MobileNet V2 encoder. Training was stopped when the network advanced 10 epochs without an improvement of at least 0.0005 in average DICE scores when assessed on the validation set. Regarding our implementation of the U-Net (Supplementary Figure 1), the key differences with the original implementation described in Ronneberger, et al. [5] for electron microscopy are the following:

1. Our input image matrix size was 256×256. The reasons for this smaller image size were practical: CT images have somewhat smaller matrix size than the electron microscopy slides. Also, smaller matrix size allowed for the U-net to require less computational resources (RAM and CPU).
2. We had 14 instead of 18 convolutional layers. Again, this is mainly due to the practical need for a smaller network, given that the input image had a smaller matrix size. For the same reasons, the network used 3 skip connections instead of 4.
3. Pooling and upsampling steps were identical and rectified Linear Units (ReLu) were used in both for activation of each convolutional layer.
4. We included a dropout step after the final convolution layer prior label prediction to reduce overfitting. 20% of data was removed during dropout. Details of the dropout are described elsewhere [6].

---

**Supplementary Table 1.** Image study characteristics for cohort

| *Characteristic* |  |
| --- | --- |
| *Study Type* | *Number of Studies* |
| CT Abd&Pel Without Contrast | 10,086 |
| CT Abd&Pel With Contrast | 19,415 |
| CT Abd&Pel With+Without Contrast | 1,923 |
| *Slice Thickness* | *Number of Scans (52,844 total scans)\** |
| 0-0.99 mm | 426 |
| 1-1.99 mm | 10248 |
| 2-2.99 mm | 5149 |
| 3-3.99 mm | 2491 |
| 4-4.99 mm | 354 |
| 5-5.99 mm | 33957 |
| > 5.99 mm | 219 |
| *Scan Information* | *mean±standard deviation* |
| Number of slices – Scan | 173±146 |
| Number of slices – Abdomen | 70±59 |
| Scan Height | 442.4±90.7 mm |
| Abdominal Compartment Height | 179.0±33.5 mm |

\*Characteristics reported for all scans that underwent abdominal classification CNN.

**Supplementary Figure 1:**
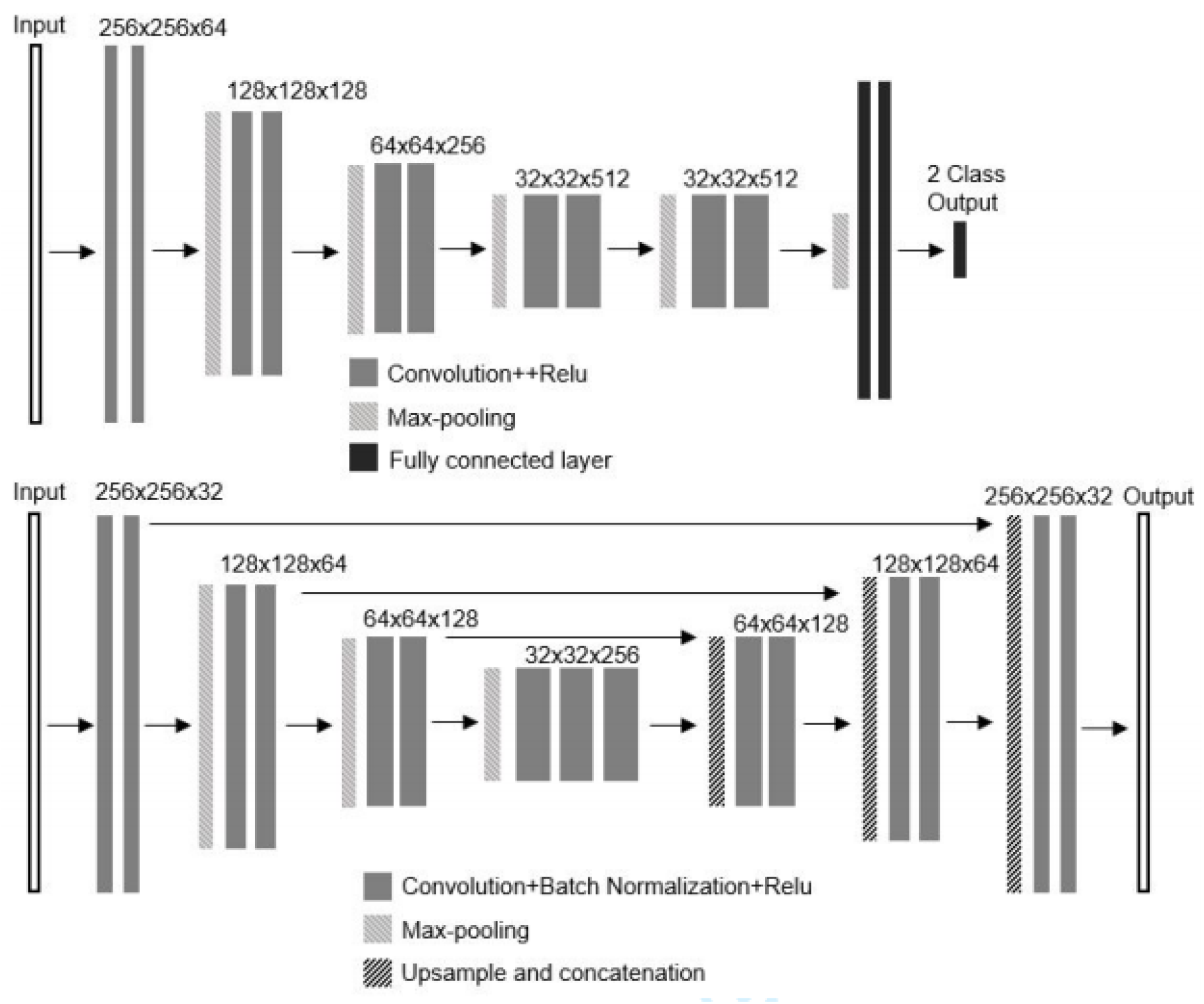
Convolutional neural networks used for A, classifying abdominal slices and B, pixel-level labeling of abdominal contour (segmentation). The neural network in A is modelled after VGG-16 and shows 5 stages of CNNs paired with two dense layers and a 2 - category output layer. The CNN in B is modelled after U-Net with 21 total layers in contracting and expanding paths with skip connections between layers of the same size. Localized feature information from the contracting path is combined with contextual information from the expanding path. The final output layer is a probability map for each pixel belonging to the quantitative imaging trait.

